# SARS-CoV-2 Viral Clearance and Evolution Varies by Extent of Immunodeficiency

**DOI:** 10.1101/2023.07.31.23293441

**Authors:** Yijia Li, Manish C. Choudhary, James Regan, Julie Boucau, Anusha Nathan, Tessa Speidel, May Yee Liew, Gregory E. Edelstein, Yumeko Kawano, Rockib Uddin, Rinki Deo, Caitlin Marino, Matthew A. Getz, Zahra Reynold, Mamadou Barry, Rebecca F. Gilbert, Dessie Tien, Shruti Sagar, Tammy D. Vyas, James P. Flynn, Sarah P. Hammond, Lewis A. Novack, Bina Choi, Manuela Cernadas, Zachary S. Wallace, Jeffrey A. Sparks, Jatin M. Vyas, Michael S. Seaman, Gaurav D. Gaiha, Mark J. Siedner, Amy K. Barczak, Jacob E. Lemieux, Jonathan Z. Li

**Affiliations:** Department of Medicine, Brigham and Women’s Hospital, Harvard Medical School, Boston, MA, USA; Department of Medicine, Massachusetts General Hospital, Harvard Medical School, Boston, MA, USA; University of Pittsburgh Medical Center, Pittsburgh, PA, USA; Ragon Institute of MGH, MIT and Harvard, Cambridge, MA, USA; Program in Health Sciences and Technology, Harvard Medical School and Massachusetts Institute of Technology, Boston, MA 02115, USA; Center for Virology and Vaccine Research, Beth Israel Deaconess Medical Center, Harvard Medical School, Boston, MA, USA; Broad Institute of MIT and Harvard, Cambridge, MA, USA

## Abstract

Despite vaccination and antiviral therapies, immunocompromised individuals are at risk for prolonged SARS-CoV-2 infection, but the immune defects that predispose to persistent COVID-19 remain incompletely understood. In this study, we performed detailed viro-immunologic analyses of a prospective cohort of participants with COVID-19. The median time to nasal viral RNA and culture clearance in the severe hematologic malignancy/transplant group (S-HT) were 72 and 40 days, respectively, which were significantly longer than clearance rates in the severe autoimmune/B-cell deficient (S-A), non-severe, and non-immunocompromised groups (P<0.001). Participants who were severely immunocompromised had greater SARS-CoV-2 evolution and a higher risk of developing antiviral treatment resistance. Both S-HT and S-A participants had diminished SARS-CoV-2-specific humoral, while only the S-HT group had reduced T cell-mediated responses. This highlights the varied risk of persistent COVID-19 across immunosuppressive conditions and suggests that suppression of both B and T cell responses results in the highest contributing risk of persistent infection.

## Introduction

Coronavirus Disease 2019 (COVID-19) vaccinations have drastically transformed the landscape of the COVID-19 pandemic by offering significant protection against infection acquisition, and severe diseases(*1, 2*), and ultimately have averted tens of millions of deaths(*3*). Unfortunately, not all individuals respond to vaccination equally well, and immunocompromised individuals can have poor vaccine responses(*4, 5*) and worse COVID-19-related outcomes(*6, 7*). Each new variant of Severe Acute Respiratory Syndrome Coronavirus-2 (SARS-CoV-2) brings risks of resistance to current treatments, particularly targeted antibody therapies(*8, 9*), resistance to vaccine-induced and naturally acquired immunity(*9, 10*), and increased transmissibility(*10*). Immunocompromised individuals have been observed to harbor detectable SARS-CoV-2 virus for longer than non-immunocompromised individuals(*11–13*). Such individuals represent a potential origin of novel SARS-CoV-2 variants as persistent infection has been associated with accelerated viral evolution(*11, 13*). However, the immunocompromised state is composed of a range of conditions and immune defects. These defects that predispose to persistent COVID-19 remain under-characterized. While there have been a number of case reports of persistent COVID-19 in immunosuppressed individuals(*11–16*) showing excessively prolonged viral shedding, persistent disease, and intra-host virological genetic diversity, there remains a need for larger scale studies with a comprehensive virologic and immunologic characterization to better elucidate the immunologic risk factors for and mechanisms of persistent infection. To this end, we hereby present a detailed longitudinal virological and immunological analysis of a cohort of immunocompromised and non-immunocompromised participants with SARS-CoV-2 infection with the goal of characterizing the virologic spectrum of persistent infection and exploring the immunologic determinants that predispose to its occurrence.

## Results

### Participant Characteristics

Fifty-six immunocompromised participants and 184 non-immunocompromised participants enrolled in the POSITIVES longitudinal cohort study were included in this analysis (*17–19*). Demographic information and key viral characteristics are shown in Table 1.

**Table 1.** Demographic and clinical information.

|  | Immunocompromised<br>(N=56) | Non-Immunocompromised<br>(N=184) | Total (N=240) | P value |
| --- | --- | --- | --- | --- |
| Sex, n (%) |  |  |  | 0.2 |
| Female | 32 (57.1%) | 126 (68.5%) | 158 (65.8%) |  |
| Male | 24 (42.9%) | 58 (31.5%) | 82 (34.2%) |  |
| Age, Median (Q1, Q3) | 55 (45, 67) | 46 (33, 59) | 49 (34-60) | 0.001 |
| Race, n (%) |  |  |  | 0.8 |
| Asian | 1 (1.8) | 10 (5.4) | 11 (4.6) |  |
| Black or AA | 5 (8.9) | 19 (10.3) | 24 (10.0) |  |
| Other/Unknown | 5 (8.9) | 16 (8.7) | 21 (8.8) |  |
| White | 45 (80.4) | 139 (75.5) | 184 (76.7) |  |
| Ethnicity |  |  |  | 0.5 |
| Hispanic or Latino | 5 (8.9) | 17 (9.2) | 22 (9.2) |  |
| Not Hispanic or Latino | 47 (83.9) | 143 (77.7) | 190 (79.2) |  |
| Other/Unknown | 4 (7.1) | 24 (13.0) | 28 (11.7) |  |
| Inpatient, n (%) | 7 (12.5) | 8 (4.3) | 15 (6.2) | 0.051 |
| Number of vaccinations,<br>median number, (Q1, Q3) | 3 (3-4) | 3 (2-3) | 3 (2-4) | <0.001 |
| mAb use, n (%) | 24* (42.9) | 10 (5.4) | 34 (14.2) | <0.001 |
| Antiviral use, n (%) | 40 (71.4) | 57 (31.0) | 97 (40.4) | <0.001 |
| Immunocompromise group, n (%) |  |  |  | <0.001 |
| S-HT | 12 (21.4) | 0 (0.0) | 12 (5.0) |  |
| S-A | 13 (23.2) | 0 (0.0) | 13 (5.4) |  |
| NS | 31 (55.4) | 0 (0.0) | 31 (12.9) |  |
| None | 0 (0.0) | 184 (100.0) | 184 (76.7) |  |
| Symptom duration,<br>median days** (Q1, Q3) | 5 (4, 7) | 4 (3, 6) | 4 (3, 6) | 0.04 |
| Variant*** |  |  |  | <0.001 |
| Delta | 3 (5.4) | 43 (23.4) | 46 (19.2) |  |
| Omicron | 48 (85.7) | 137 (74.5) | 185 (77.1) |  |
| Other/Unknown | 5 (8.9) | 4 (2.2) | 9 (3.8) |  |

Q1 and Q3, quartile 1 and quartile 3; AA, African American; mAb, monoclonal antibody; S-HT, severe with malignant hematology or transplant history; S-A, severe autoimmune/B-cell deficient; NS, non-severe immunocompromising condition.
\*, four participants received Mab after blood draws.
\*\*, Symptom duration indicates the duration between symptom onset (patient report or first positive test if asymptomatic screening) and first nasal swab collected by the study group.
\*\*\*, Variant information was obtained by either Spike or whole genome sequencing or by epidemiological information (time period when the participant was infected).

Immunocompromised participants were significantly older than non-immunocompromised controls (median 55 vs 46 years, P=0.001), and more likely to receive monoclonal antibody (mAb) and/or antiviral treatment against SARS-CoV-2. The two groups had comparable sex, race, and ethnicity profiles and a similar median time from symptom onset/first positive COVID-19 test to enrollment (5 vs 4 days). We further subdivided immunocompromised participants into the severe hematologic-oncology/transplant (S-HT, n=12), severe autoimmune/B-cell deficient (S-A, n=13) and non-severe (NS, n=31) groups (refer to Supplementary Tables 1 and 2 for detailed categorization). Three participants died due to severe COVID-19 or COVID-19 related complications, all of whom were in the immunocompromised sub-group (S-HT and S-A).

### Delayed Viral Clearance in Hematologic Oncology and Transplant Participants

We first aimed to characterize viral dynamics in the upper respiratory tract in participants with different categories of immunocompromising conditions. Immunocompromised and non-immunocompromised participants had similar peak vRNA levels (5.1, 5.1, 4.9, and 5.7 log_10_ SARS-CoV-2 copies/ml in S-HT, S-A, NS and non-immunocompromised groups, P=0.5). However, the rates of nasal vRNA decay were significantly different between the immunocompromising categories, with the S-HT group demonstrating significantly slower viral clearance compared to other groups (Fig. 1a and 1b). Median time to nasal vRNA clearance in the S-HT group was 72 days (95% confidence interval [CI] 5, not available [NA]) compared to 7 (6, NA) days for the S-A, 11 (8, 16) days for the NS and 11 (10, 12) days for the non-immunocompromised group (Fig. 1b, Log-rank P=0.002). Similarly, the S-HT group experienced a significant delay in the clearance of culturable virus (Fig. 1c and 1d). Median time to viral culture clearance in the S-HT group was 40 days (95%CI 5, NA) compared to 6.5 (5, NA) days for the S-A group, 6 days (5, 7) for the NS group and 7 days (6, 7) for the non-immunocompromised group (Fig. 1d, log-rank P<0.001, Fig. 1d). After 30 days from symptom onset or first positive test, 50%, 15%, and 6.5% of participants from S-HT, S-A and NS groups had detectable vRNA, compared to 0% in non-immunocompromised group (P<0.0001, Supplementary Fig. 1a). In addition, 50% of S-HT and 8.3% of S-A participants still had culturable virus, compared to 0% in the NS and non-immunocompromised groups after 30 days (P<0.0001, Supplementary Fig. 1b). Compared to the non-immunocompromised group, the S-HT group was significantly associated with delayed vRNA clearance (adjusted hazard ratio [aHR] for viral clearance, 0.32, 95%CI 0.12-0.83, P=0.02) and culturable virus clearance (aHR 0. 27 for viral clearance, 95% CI 0.12-0.63, P=0.002), after adjusting for demographics, number of vaccinations and antiviral use (Table 2).

**Figure 1.**
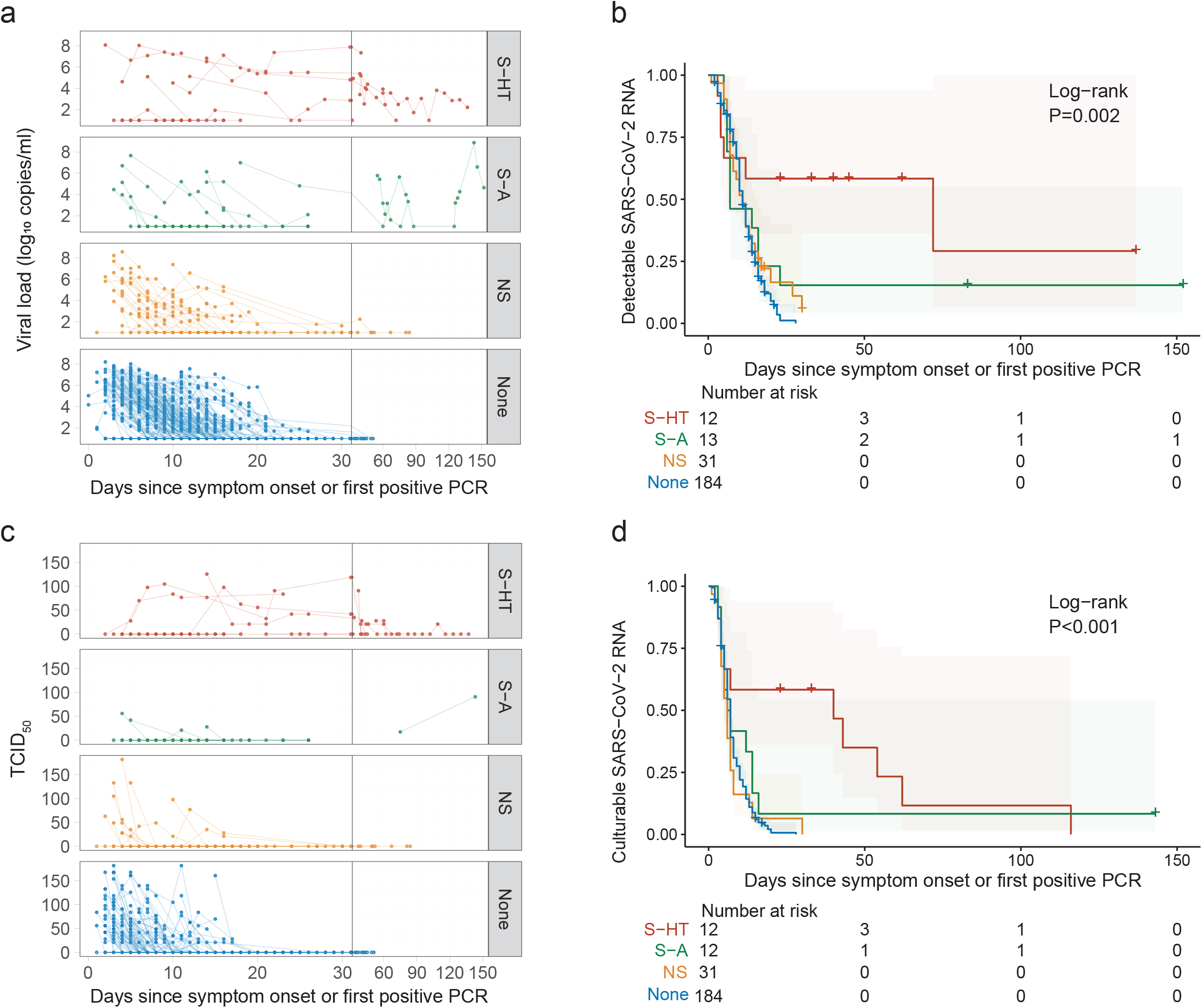
Kinetics of SARS-CoV-2 viral RNA and culturable virus among different immunocompromised groups. **a**, Upper respiratory viral load decay. Lower level of quantification (LLOQ) is 10 copies/ml. **b**, Kaplan-Meier estimates of upper respiratory viral clearance (viral load below LLOQ). **c**, Upper respiratory culturable virus dynamics (50% Tissue Culture Infectious Dose [TCID_50_)). **d**, Kaplan-Meier estimates of upper respiratory culturable virus clearance.

**Table 2.** Association between immunocompromise groups and SARS-CoV-2 viral decay.

| Hazard ratio for SARS-CoV-2 viral RNA clearance |  |  |  |  |
| --- | --- | --- | --- | --- |
| Group | HR (95% CI) | P | aHR (95% CI) | P |
| None (Reference) |  |  |  |  |
| S-HT | 0.22 (0.09-0.52) | 0.0005 | 0.32 (0.12-0.83) | 0.02 |
| S-A | 0.64 (0.34-1.22) | 0.2 | 0.74 (0.33-1.64) | 0.5 |
| NS | 0.82 (0.54-1.24) | 0.4 | 0.92 (0.58-1.46) | 0.7 |
| Hazard ratio for SARS-CoV-2 culturable virus clearance |  |  |  |  |
| Group | HR (95% CI) | P | aHR (95% CI) | P |
| None (Reference) |  |  |  |  |
| S-HT | 0.23 (0.11-0.48) | 8.7E-5 | 0.27 (0.12-0.63) | 0.002 |
| S-A | 0.54 (0.28-1.03) | 0.06 | 0.53 (0.26-1.11) | 0.09 |
| NS | 1.12 (0.76-1.66) | 0.6 | 1.35 (0.89-2.06) | 0.2 |

HR, hazard ratio; aHR, adjusted hazard ratio; CI, confidence interval.
Age, sex at birth, race, ethnicity, number of vaccinations, monoclonal use, and antiviral use were adjusted for in the multivariate models.

### Increased Viral Evolution and Genetic Diversity in Immunocompromised Participants

We used gene-specific next-generation sequencing approach to quantify the number of unique intra-host single-nucleotide variants (iSNVs) in the S gene present at >3% frequency within each sample. This analysis was limited to participants with a viral genome available both at baseline and at least one follow-up timepoint. Severely immunocompromised (S-HT and S-A) participants harbored a greater number of iSNVs over time compared to NS and non-immunocompromised group participants, although these comparisons did not reach statistic significance (Fig. 2a). To evaluate viral diversity, we calculated the average pairwise distance both at the nucleotide and at the amino-acid level. Nucleotide average pairwise distance was significantly higher in the severe immunocompromised (S-HT and S-A) group compared to either non-severe (4.0E-4 vs 8.2E-5, P<0.001 Dunn’s test with Benjamini-Hochberg adjustment) or non-immunocompromised groups (4.0E-4 vs 2.3E-5, P<0.001, Fig. 2b). Similar results were obtained when the average pairwise distances were calculated for amino acids (Supplementary Fig. 2a). Among participants with longitudinal sequences, 39% of the participants in immunocompromised group versus 12% in the non-immunocompromised group had evidence of viral nucleotide changes (Fisher’s exact P<0.001, Fig. 2c). These nucleotide changes were distributed across the entire length of the S gene (Fig. 2d).

**Figure 2.**
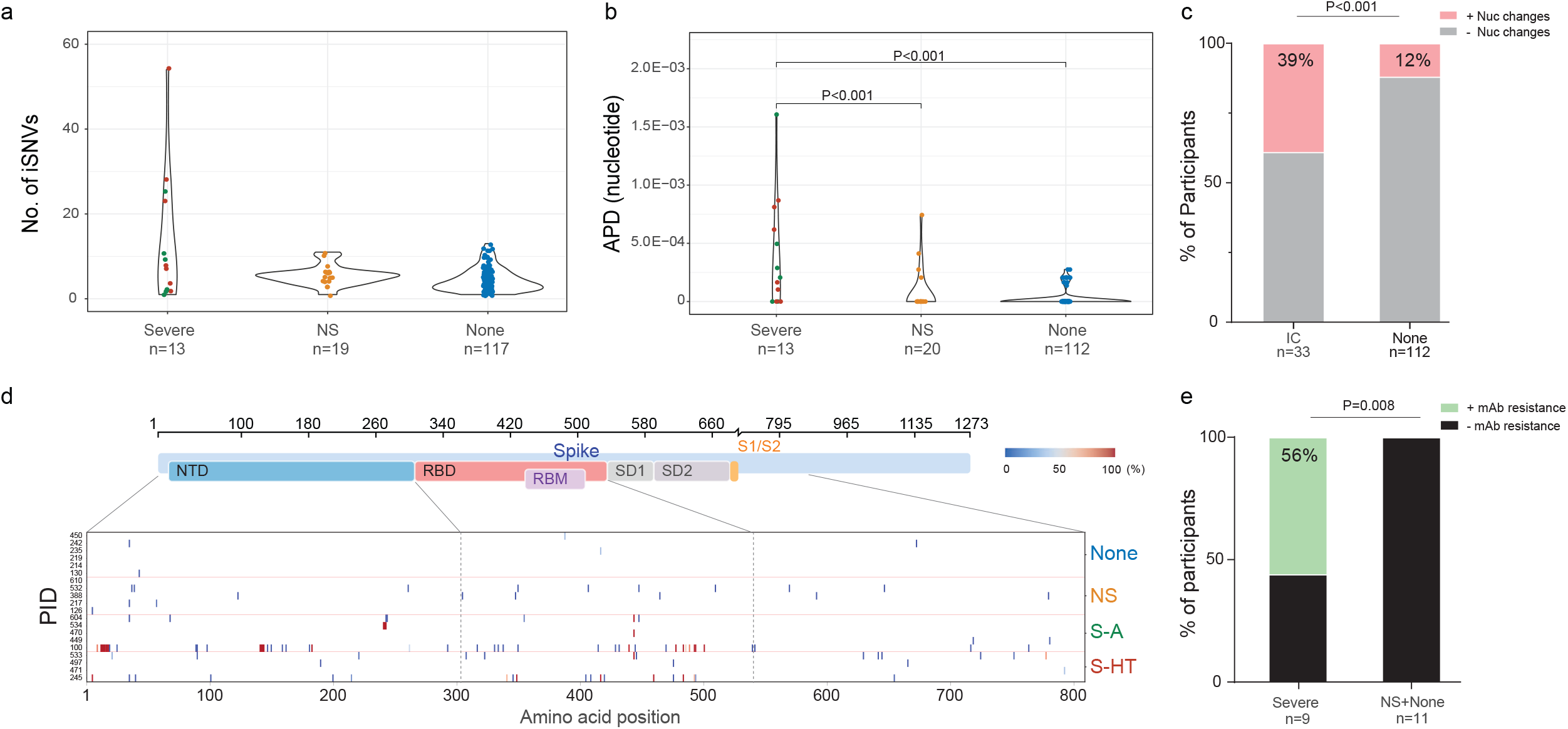
SARS-CoV-2 intra-host mutations among different immunocompromise groups. **a**, Number of intra-host single-nucleotide variants (iSNVs) among severe (S-HT in red and S-A in green), non-severe immunocompromised and non-immunocompromised (None) groups. **b,** Nucleotide average pairwise distance (APD) among severe (S-HT in red and S-A in green), non-severe immunocompromised (NS) and non-immunocompromised (None) groups. **c**, Participants with any nucleotide changes during follow-up. **d**, Heat map showing distribution of Spike polymorphisms from participants receiving mAb treatment. Each row represents one participant, while x axis shows amino acid positions in the Spike gene. Different domains of Spike are shown at the top. Colors indicate frequency of polymorphisms, with blue indicating the lowest value and red indicating the highest value in the scale. Participants in different study groups are separated by a red horizontal line. **e**, Proportion of mAb resistance emergence amongst those treated with mAbs, categorized by those with severe or non-severe/no immunosuppression. Comparison of iSNV and APD between groups were done using using Dunn’s test with Benjamini-Hochberg P value adjustment. Fisher’s exact test was used to calculate significance between participants with and without viral evolution and further, in participants with and without mAb treatment specific resistance mutations. Only significant P values are shown. NTD, N-terminal domain; RBD, receptor binding domain; RBM, receptor binding motif; S1, subunit 1; S2, subunit 2.

### Increased Risk of Treatment-Emergent Resistance to Anti-SARS-CoV-2 Monoclonal Antibody Therapy

Deep sequencing analysis of Spike gene was carried out to evaluate the dynamics of mutation emergence in the presence of mAb treatment as earlier reports have shown evidence of mAb resistance emergence both in immunocompromised and non-immunocompromised participants(*13, 20–29*). In total, 34 participants across different study groups received mAb therapy, 10 in S-HT, 9 in S-A, 5 in NS, and 10 in non-immunocompromised groups (Details in Supplementary Table 3). Of these, we were able to evaluate the risk of resistance emergence in a subset of participants for whom sequences were available at both baseline and at least one follow-up time point. Five out of nine (56%) severely immunocompromised participants (S-HT and S-A) developed mAb-specific resistance mutations (Supplementary Fig. 2b). This was a significantly higher rate than that found in the non-severe or non-immunocompromised groups (0/11 [0%] combined, Fisher’s exact P=0.008) (Fig 2e).

### Suboptimal Humoral Response in Severely Immunocompromised Participants

We next characterized the antibody response in immunocompromised and non-immunocompromised participants. In participants whose serum sample was available (n=94), including those who had previously received monoclonal antibodies, we found no significant difference in nAb titers between the different immunocompromised and non-immunocompromised groups at early and later sampling time points, although this analysis may have been limited due to individuals who received anti-SARS-CoV-2 monoclonal antibody infusion before sample collection (either therapeutic or pre-exposure prophylaxis) (Fig. 3a and 3b). The non-immunocompromised group had a significant increase in anti-ancestral and anti-variant spike nAb levels during follow-up, whereas we did not observe a significant increase in antibody levels in the immunocompromised group (Fig. 3a and 3b). However, after excluding individuals with exposure to mAb therapies, the severe immunocompromised group showed no significant changes in either anti-ancestral or anti-variant Spike nAb levels, while the non-severe immunocompromised group demonstrated a moderate increase and the non-immunocompromised group demonstrated the greatest increase in nAb levels, plateauing at day 25-30 (Fig. 3c and d).

**Figure 3.**
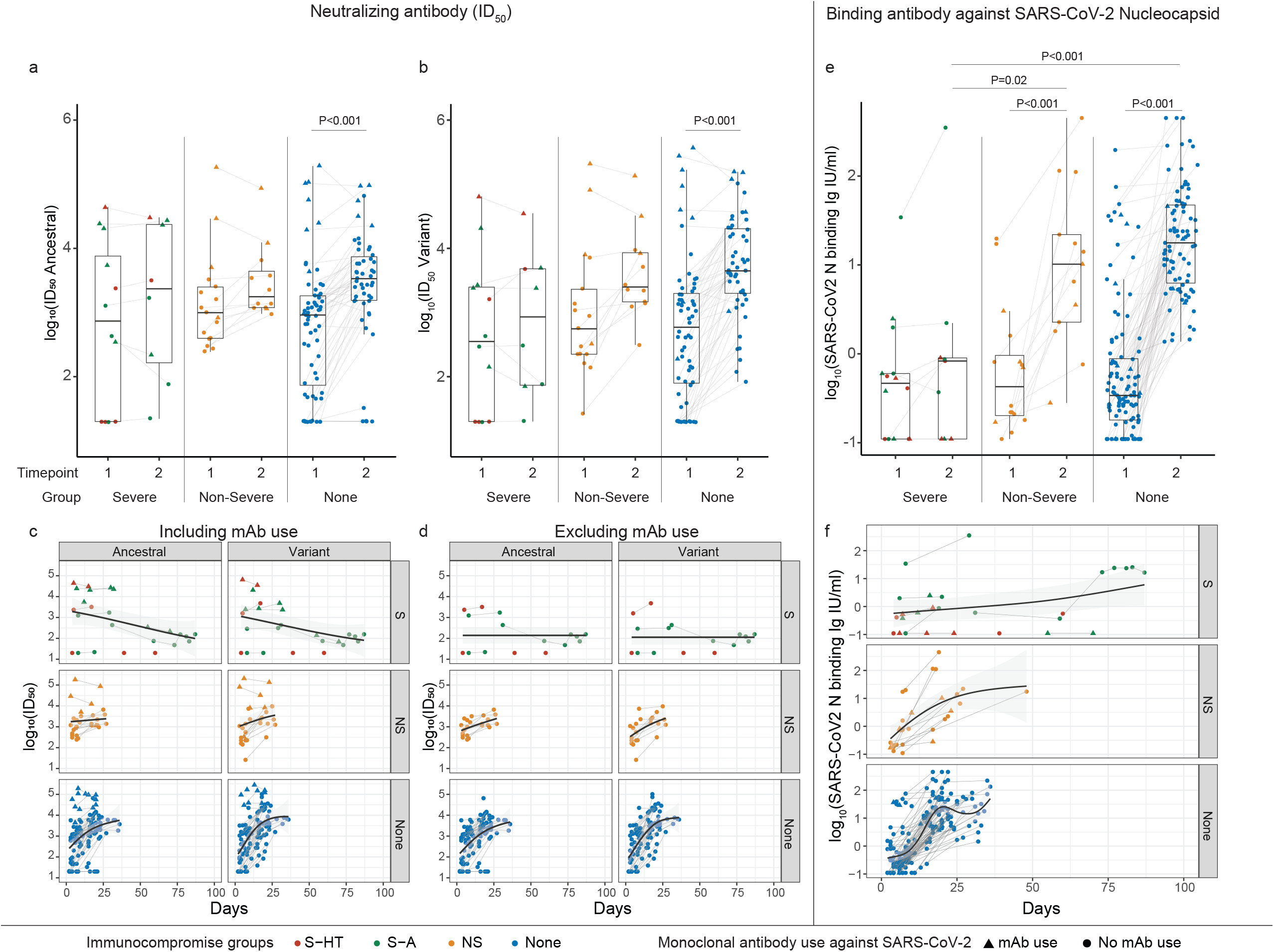
Neutralizing antibody (nAb) and Nucleocapsid binding antibody levels among different immunocompromised groups. **a**, nAb levels (50% inhibitory dilution [ID_50_]) against ancestral Spike protein. **b**, nAb levels against variant-specific Spike protein. **c** and **d**, Longitudinal trajectory of nAb in different immunocompromise groups, including (**c**) or excluding (**d**) monoclonal antibody use. **e**, binding antibody levels against Nucleocapsid protein. **f**, Longitudinal trajectory of binding antibody in different immunocompromise groups. Comparison between different immunocompromise groups at the same time point was performed using Dunn’s test with Benjamini-Hochberg P value adjustment. Comparison of longitudinal antibody changes for participants with two blood draws was performed using the pairwise Wilcoxon rank sum test with Benjamini-Hochberg P value adjustment. Only significant P values were shown. Tukey boxplot was used to summarize antibody levels. Generalized additive model was used to evaluate the trend of antibody development with 95% confidence intervals in the shaded area. Lines between two timepoints indicate the same participants with two blood draws. S-HT, severe hematologic-oncology/transplant; S-A, severe autoimmune/B-cell deficient; NS, non-severe. Severe group included both S-HT and S-A as they had comparable antibody levels at multiple time points.

In a generalized estimating equation (GEE) model adjusted for factors associated with nAb levels, severely immunocompromised participants were only able to mount 0.18-fold (approximately 5-fold lower) of the anti-ancestral Spike antibody (95% CI 0.05-0.60) and 0.08-fold (approximately 12-fold lower) of the anti-variant Spike antibody (95% CI 0.03-0.22) compared to non-immunocompromised participants (Supplementary Fig. 3a, b). Non-severe immunocompromised status was not associated with significant differences in nAb changes from acute to post-infection, compared to non-immunocompromised group. In the whole cohort, each dose of vaccinations prior to SARS-CoV-2 infection was associated with 1.70-fold (95% CI 1.25 to 2.30) and 1.35-fold (95% CI 1.01 to 1.79) increase in anti-ancestral and anti-variant Spike antibody levels, respectively (Supplementary Fig. 3ab).

We also evaluated binding antibody against nucleocapsid protein because this assay is not affected by monoclonal antibody use. Similar to nAb, individuals in the S-HT and S-A sub-groups had significantly blunted increases in nucleocapsid binding Ab development from acute to post-infection, and significantly lower binding Ab levels compared to NS and non-immunocompromised groups (Fig. 3e). Longitudinally, binding Ab levels in NS and non-immunocompromised groups plateaued around day 20-25 at a level of 1-1.5 log_10_ IU/ml, while S-HT/S-A groups exhibited delayed development to a level below 1 log_10_ IU/ml after day 50 (Fig. 3f).

### T Cell Responses

In a recent study, Apostolidis et al. demonstrated elevated spike-specific CD8^+^ T cell responses in COVID-19 mRNA vaccinated participants with multiple sclerosis receiving anti-CD20 treatment compared to healthy controls(*30*). However, it remains largely unknown if different types and levels of immunosuppression are associated with a similar T cell immunophenotype. To this end, we profiled the T cell effector function using enzyme-linked immunosorbent spot (ELISpot) and antigen-specific proliferation assay for a selected group of participants based on sample availability. The non-immunocompromised group had lower interferon gamma (IFN-γ) producing units per million cells upon stimulation at both blood draws (acute infection 0-14 days, and post-acute 15-60 days after symptom onset or first positive COVID-19 test) compared to both NS and S-A groups in response to both ancestral and variant-specific Spike peptide pools (Fig. 4a). Individuals in the S-A group tended to have the highest levels of CD4+ and CD8^+^ T cell proliferation upon spike peptide pool stimulation, especially compared to the S-HT and non-immunocompromised individuals (Fig. 4bc and Supplementary Fig. 4). In longitudinal analysis, the S-HT group showed poor functional CD4^+^ and CD8^+^ T cell proliferation, despite a comparable IFN-γ secretion level compared to the non-immunocompromised group (Fig. 4d). In contrast, the S-A group showed robust T cell proliferation over time, in response to both ancestral and variant-specific spike peptide pools compared to all other groups, suggestive of either an antigen-stimulated compensatory effect in the setting of B cell deficiency or a medication-related T cell priming.

**Figure 4.**
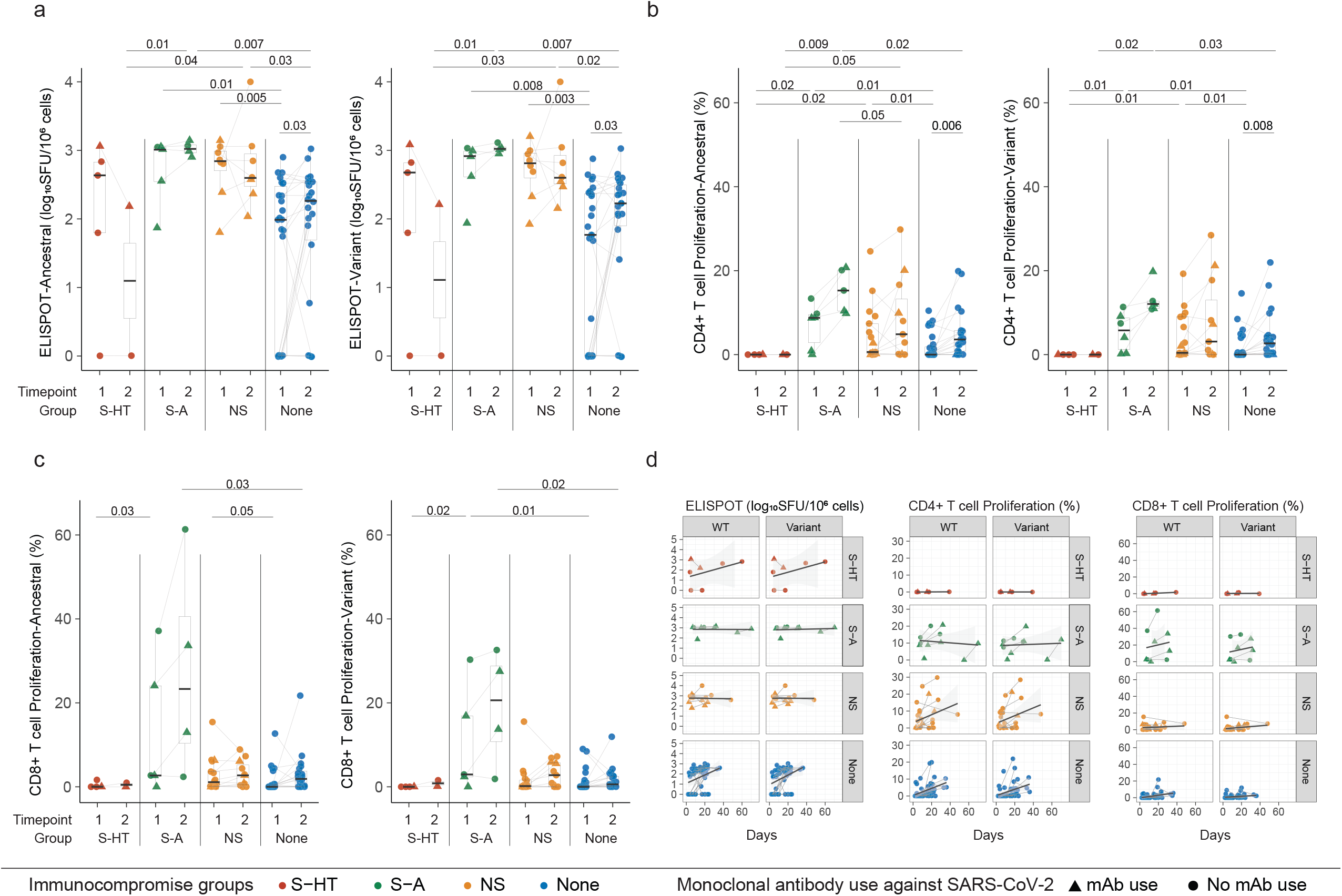
Spike-specific T levels among different immunocompromised groups. **a**, enzyme-linked immunosorbent spot (ELIspot) assays using peptide pools derived from ancestral and variant-specific Spike protein. **b, c**, CD4^+^ T cell (b) and CD8^+^ T cell proliferation (c) upon stimulation of ancestral- and variant-specific Spike peptide pools. **d**, Longitudinal trajectory of Spike-specific T levels in different immunocompromise groups. Comparison between different immunocompromise groups at the same time point was performed using Dunn’s test with Benjamini-Hochberg P value adjustment. Comparison of longitudinal antibody changes for participants with two blood draws was performed using the pairwise Wilcoxon rank sum test with Benjamini-Hochberg P value adjustment. Only significant P values were shown. Tukey boxplot was used to summarize antibody levels. Generalized additive model was used to evaluate the trend of antibody development with 95% confidence intervals in the shade area. Lines between two timepoints indicate the same participants with two blood draws.

## Discussion

Understanding viral and immune control characteristics of COVID-19 infection is crucial to our ability to care for immunocompromised individuals at greatest risk of persistent and severe infection. Moreover, it can guide public health interventions and shed light on vaccine development that protects immunocompromised individuals. In this study, we performed an in-depth virologic and immunologic evaluation of a cohort of immunocompromised and non-immunocompromised individuals. We demonstrated a hierarchy of immunocompromised conditions that increase the risk of delayed viral clearance and SARS-CoV-2 evolution.

Furthermore, we identified the varied risk of persistent COVID-19 across immunosuppressive conditions and suppression of both B and T cell responses in those at the highest risk of persistent infection. Specifically, we found that individuals with a history of hematological malignancy and organ transplant demonstrated the greatest delay in viral clearance that may be mediated by suppression of both the B and T cell responses. In contrast, those with B cell immunodeficiency had an intermediate risk of chronic infection in the setting of an immune response showing compensatory heightened levels of SARS-CoV-2-specific T cell function.

Our cohort study confirmed findings from prior case reports and case series. In a review by Dioverti et al., the authors summarized cases of persistent COVID-19 lasting from approximately one month to one year(*31*). These included a spectrum of immunocompromised hosts from individuals with solid organ transplants, hematological malignancy(*14, 20, 22, 23, 25, 29, 32–36*), and autoimmune diseases receiving immunosuppressive therapies. However, immunocompromise is a broad spectrum and, until now it has not been clear which immunosuppressive conditions represent the greatest risk for persistent infection. The results of our study provide critical insight as to the hierarchy of risk for delayed SARS-CoV-2 RNA and culture clearance, and viral evolution. Specifically, we found that demonstrated that individuals with solid organ transplant and hematological malignancy are associated with the longest period of viral RNA and culturable viral shedding, followed by severely immunocompromised participants with autoimmune conditions receiving B-cell depleted therapy and/or those with B cell deficiency. Participants with mild non-severe immunocompromise, such as those with autoimmune diseases receiving anti-tumor necrosis factor (TNF) treatment, had similar viral shedding dynamics to non-immunocompromised participants.

There have been several prior case reports of immunosuppressed individuals with chronic COVID-19 and accumulation of viral polymorphisms and drug resistance mutations(*13, 14, 21, 27, 34*), but there has been little in the way of more systematic evaluation of viral evolution. In this study, we used S gene-specific deep sequencing to assess longitudinal viral evolution and diversity. Our results show that severe immunosuppression is associated with increased viral evolution and diversification. In addition, severe immunocompromised hosts had a greater risk of developing treatment-emergent resistance mutations to mAb therapy when compared to non-immunocompromised group participants. These findings highlight the potential for immunocompromised individuals to serve as a source for SARS-CoV-2 evolution and drug resistance, consistent with isolated reports of immunocompromised hosts implicated in the emergence of highly mutated SARS-CoV-2 variants(*13, 15, 16, 24, 28, 37–40*). It should be noted though that even within the category of severe immunocompromise, participants demonstrated a range of viral diversification and evolution patterns, and additional studies are needed to fully assess the drivers of accelerated viral evolution.

Another highlight of our study is our use of in-depth analysis of B and T cell responses, including SARS-CoV-2-specific neutralizing and binding antibody levels, as well as ELISpot and T cell proliferation studies. We noted lower levels of neutralizing antibody against both ancestral virus and variant virus in the severe immunocompromised group (less than 10-20% compared to non-immunocompromised individuals), after adjusting for vaccination doses, monoclonal antibody use, and demographic confounders. Each additional dose of vaccination is associated with an approximately 1.5-fold increase in antibody levels in the whole cohort, underscoring the importance of adherence to COVID-19 vaccination recommendations. However, there is evidence that non-B-cell immunity may be sufficient for the clearance of SARS-CoV-2. Early in the pandemic, cases were reported of individuals with X-linked agammaglobulinemia who developed COVID-19 pneumonia, but subsequently recovered despite a lack of SARS-CoV-2-specific immunoglobulins(*41*). In our study, the risk of chronic infection was highest in S-HT participants. This group of participants was found to have both suboptimal humoral and cell-mediated immune responses. The “near-normal” level of effector ELISpot responses in S-HT individuals, compared to the non-immunocompromised group, is the likely the result of exposure to high levels of SARS-CoV-2 antigen, while the reduced proliferation demonstrates compromised functionality. In contrast, the S-A participants had an even more robust SARS-CoV-2-specific proliferative T-cell responses than the non-immunocompromised group, indicating increased levels of functional SARS-CoV-2-specific CD8^+^ T cell responses, which was associated with an intermediate risk of chronic infection. These results align with some intriguing reports that individuals receiving anti-CD20 treatment may demonstrate a stronger T-cell responses, in particular more robust activation-induced marker-positive CD8^+^ T cell responses(*30*). We found that both CD8^+^ T cell response and CD4^+^ T-cell responses, including proliferation in response to both ancestral and variant-specific spike peptides, were more pronounced in S-A group compared to other non-immunocompromised groups. Together with the results from Apostolidis et al.(*30*), our results raise the question of whether individuals with B cell deficiencies may have a lower risk of persistent SARS-CoV-2 infection due to preserved T cell function, either as a compensatory mechanism or T cell priming by certain B cell-depleting therapies.

There are several limitations to this study. We included a relatively small number of individuals with malignant hematological conditions or transplant history, and we were able to obtain blood samples from only a subset of the participants to characterize their humoral and T-cell immunity. Larger studies are needed to provide greater precision as to extent of immune defect or immunosuppressive medication that may place patients at the greatest risk of chronic infection and viral evolution. We also did not analyze markers reflecting innate immunity, including soluble inflammatory markers and monocyte phenotypes. Our study focused on virologic and immunologic responses after COVID-19 and it is unclear how these may contribute to persistence and severity of symptoms. Furthermore, we only evaluated Spike-specific humoral and cellular immunity, while immunity targeting other structural or non-structural proteins has been shown to alter disease course(*42, 43*).

In conclusion, in this prospective cohort of well-characterized individuals with acute COVID-19, we demonstrated a correlation between a hierarchy of immunocompromised conditions and SARS-CoV-2 viral shedding, viral evolution, and adaptive immunity. Our results highlight the finding that the risk of chronic SARS-CoV-2 infection is not uniform across immunosuppressive conditions and provide clarity on which immunosuppression conditions predispose to delayed SARS-CoV-2 RNA and culture clearance, and viral evolution. The high risk populations identified in this study may benefit from targeted public health and additional therapeutic interventions. In addition, the results provide further insights on the humoral and cell-mediated immune correlates of viral clearance, which is crucial for the development of improved vaccines and future therapies.

## Materials and Methods

### Participant Enrollment and Sample Collection

We enrolled participants with a positive COVID-19 test in the Mass General Brigham Medical HealthCare System as part of the POSt-VaccInaTIon Viral CharactEristics Study (POSITIVES) (*17, 19*) in addition to one immunocompromised participant from our previous study. Each participant’s medical record was reviewed for demographic data, immunosuppression status, and COVID-19 treatment history by board-certified clinicians. For the POSITIVES study, participants self-collected anterior nasal swabs every 2-3 days to a total of 6 samples over 2 weeks. Participants with immunocompromised conditions were offered to collect additional swabs when possible and were followed until they had two consecutive negative PCR tests. For this one immunocompromised participants reported previously(*13*), nasopharyngeal swab was collected by healthcare providers. In a subset of participants who agreed to provide blood sample, the first blood draw was done generally before day 15 of symptom onset (acute phase) or first positive PCR or antigen test for COVID-19, and second blood draw between 15-60 days after (post-acute phase) (Supplementary Fig. 5). This study was approved by Mass General Brigham Institutional Review Board and all participants have signed informed consent upon entry to the study.

### Categorization for Immunocompromised Conditions

Immunocompromised participants were further categorized into the following groups: severe immunocompromised participants, which were further categorized into severe-hematological malignancy/transplant patients (S-HT), severe autoimmune patients (S-A, participants with autoimmune condition receiving B-cell targeting agents or B cell deficiency); and non-severe immunocompromised participants (NS). This categorization was based on a recent cohort study which demonstrated a hierarchy of antibody response to COVID-19 vaccinations in different medical conditions(*4, 44*). Detailed classification criteria were listed in Supplementary Table 1.

### SARS-CoV-2 Viral Load Assay

SARS-CoV-2 viral RNA (vRNA) were quantified as described previously(*45*). Briefly, virions were pelleted from nasal swab fluid by centrifugation at 21,000g for 2 hours at 4°C. Trizol-LS Reagent (Thermo fisher Scientific, Waltham, MA) was added to the pellet, vortexed, and incubated on ice for 10 minutes. Chloroform was added and the solution was vortexed before centrifugation at 21,000g for 15 minutes at 4°C. RNA was isolated from the aqueous layer by isopropanol precipitation and eluted in DEPC-Treated water (Thermo fisher Scientific, Waltham, MA). SARS-CoV-2 RNA copies were quantified with an in-house viral load assay using the CDC 2019-nCoV_N1 primer and probe set (Integrated DNA Technologies, Coralville, IA). The efficiency of the RNA extraction and RT-qPCR amplification was evaluated by quantifying the RCAS RNA recovered from each sample and the two N1 controls. The importin-8 (IPO8) human housekeeping gene was also amplified and evaluated as a measure of sample collection quality. Samples were run in triplicate wells for N1, and in duplicate wells for RCAS and IPO8.

### SARS-CoV-2 Viral Culture Assay

Viral culture was performed as previously reported(*17*). Vero-E6 cells (ATCC, Manassas, VA) maintained in DMEM (Corning, Corning, NY) supplemented with HEPES (Corning, Corning, NY), 1X Penicillin (100 IU/mL)/Streptomycin (100 μg/ml) (Corning, Corning, NY), 1X Glutamine (Glutamax, Thermo fisher Scientific, Waltham, MA) and 10% FBS (MilliporeSigma, Burlington, MA) were plated 16-20 hours before infection. Each sample consisting of nasal swab fluid was thawed on ice and filtered through a Spin-X 0.45 um filter (Corning, Corning, NY) at 10,000 xg for 5 minutes. Before infection the media was changed to DMEM supplemented with HEPES, 1X Antibiotic/Antimycotic (Thermo Fisher, Waltham, MA), 1X Glutamine, 2% FBS and 5 μg/mL of polybrene (Santa Cruz Biotechnologies, Dallas, TX). Each filtered sample was then used to inoculate Vero-E6 cells by spinfection (2,000x g for 1 hour at 37C). Each condition was plated in quadruplicate wells in 1:5 dilutions across half the plate. The plates were observed at 7-days post-infection using a light microscope to check for cytopathogenic effect (CPE) and a median tissue culture infectious dose (TCID_50_) was calculated for each sample.

### Neutralizing Antibody Responses

Neutralizing activity against SARS-CoV-2 pseudovirus was measured using a single-round infection assay in 293T/ACE2 target cells(*18*). Pseudotyped virus particles were produced in 293T/17 cells (ATCC, Manassas, VA) by co-transfection of plasmids encoding codon-optimized full-length Spike (ancestral-D614G, Delta, Omicron-BA.1, Omicron-BA.2, Omicron-BA.4/5), packaging plasmid pCMV ΔR8.2, and luciferase reporter plasmid pHR’ CMV-Luc. Packaging and luciferase plasmids were kindly provided by Dr. Barney Graham (NIH, Vaccine Research Center). The 293T cell line stably overexpressing the human ACE2 cell surface receptor protein was kindly provided by Drs. Michael Farzan and Huihui Ma (The Scripps Research Institute).

For neutralization assays, serial dilutions of patient sera were performed in duplicate followed by addition of pseudovirus. Pooled serum samples from convalescent COVID-19 patients or pre-pandemic normal healthy serum (NHS) were used as positive and negative controls, respectively. Plates were incubated for 1 hour at 37℃ followed by addition of 293/ACE2 target cells (1x10^4^/well). Wells containing cells + pseudovirus (without sample) or cells alone acted as positive and negative infection controls, respectively. Assays were harvested on day 3 using Promega BrightGlo luciferase reagent and luminescence detected with a Promega GloMax luminometer (Promega, Madison, WI). Titers are reported as the dilution of serum that inhibited 50% virus infection (ID_50_ titer). Pseudovirus-based neutralization assays were conducted using ancestral Spike protein, as well as Delta- and Omicron- (BA.1, BA.2, or BA.4/5) Spike. Anti-variant neutralizing antibody level (nAb) was determined based on the viral strain each participant was infected with (either by sequencing or in small proportion, imputed by time of infection when specific strain was prevalent).

### Nucleocapsid binding antibody assay

Binding antibody against Nucleocapsid protein was measured using Coronavirus Ig Total Human 11-Plex ProcartaPlex™ Panel (Thermo fisher Scientific, Waltham, MA) according to the manufacturer’s instruction.

### T Cell Enzyme-linked immunosorbent spot (ELISpot) Assay

Interferon (IFN)-γ ELISpot assay was reported in our previous study and were performed according to the manufacturer’s instructions (Mabtech, Cincinnati, OH)(*46*). Briefly, peripheral blood mononuclear cells (PBMCs) were incubated with SARS-CoV-2 peptide pools (MGH Peptide Core) at a final concentration of 0.5μg/ ml for 16–18h (100,000-200,000 cells per test). Anti-CD3 (Clone OKT3, Biolegend, 0.5μg/mL, San Diego, CA) and anti-CD28 Ab (Clone CD28.2, Biolegend, 0.5μg/mL, San Diego, CA) were used as positive controls. To quantify antigen-specific responses, mean spots of the DMSO negative control wells were subtracted from the positive wells. The results were expressed as spot-forming units (SFU) per 10^6^ PBMCs. Responses were considered positive if the results were >5 SFU/10^6^ PBMCs following control subtraction. If negative DMSO control wells had >30 SFU/10^6^ PBMCs or if positive control wells (anti-CD3/anti-CD28 stimulation) did not have >1000 spot-forming units, the results were deemed invalid and excluded from further analysis.

### T Cell Proliferation Assay

T Cell proliferation assay was reported previously(*46*). Briefly, PBMCs were incubated in PBS with 0.5 uM carboxyfluorescein succinimidyl ester (CFSE; Life Technologies, Carlsbad, CA) or CellTrace™ Far Red (CTFR, Invitrogen, Waltham, MA) at 37°C for 20 min. Then they were washed and resuspended at 0.5-1 × 10^6^/mL and plated into 96-well U-bottom plates (Corning) in 200 µL of. media. Peptide pools were added at a final concentration of 0.5 µg/mL, followed by incubation at 37°C, 5% CO_2_ for six days. The PBMC staining antibody panels are in Supplementary Materials. Cells were washed and fixed in 2% paraformaldehyde prior to flow cytometric analysis on a BD LSR II (BD Biosciences, Franklin Lakes, NJ). A positive proliferation response was defined as a percentage of CD3^+^CD4^+^ or CD3^+^CD8^+^ CFSE^low^ or CTFR^low^ cells with at least 1.5x greater than the highest of two negative-control wells and greater than 0.2% CFSE^low^ or CTFR^low^ cells in magnitude following background subtraction.

### SARS-CoV-2 S-gene Sequencing

SARS-CoV-2 viral RNA (vRNA) isolation as described previously(*45*). RNA was converted to cDNA using Superscript IV reverse transcriptase (Invitrogen, Waltham, MA) as per manufacturer’s instructions). Spike (S) gene amplification was performed using a nested PCR strategy with in-house designed primer sets targeting codons 1–814 of the Spike as previously described(*28*). Further, PCR products from different individuals were pooled, and Illumina library construction was performed using the Nextera XT library prep kit (Illumina). Sequencing was performed on the Illumina MiSeq platform and deep sequencing data analysis was carried out using the Stanford Coronavirus Antiviral & Resistance Database (CoVDB) platform (https://covdb.stanford.edu/sierra/sars2/by-reads/?cutoff=0.01&mixrate=0.01)(47). Input FASTQ sequence alignment with Wuhan-Hu-1 reference was done using MiniMap2 version 2.22 in CodFreq pipeline (https://github.com/hivdb/codfreq). The output of MiniMap2, an aligned SAM file, is converted to a CodFreq file by an in-house written Python script using a PySam library (version: 0.18.0) and further analyzed with the CoVDB. PCR and sequencing runs were performed once with the appropriate positive and negative controls. For S gene analysis, amino acid variants were then called at the codon level using perl code and used for resistance interpretation with a 1% limit of detection. The accuracy of the deep sequencing platforms was evaluated with a control library of clonal SARS-CoV-2 sequences mixed at known concentrations as described previously(*48*). Mutations detected by next-generation sequencing at <20% of the viral population were labelled as ‘low-frequency’ variants as they would largely be missed by traditional Sanger sequencing. A minimum average of 500x sequencing coverage per sample was required for variant calling. SARS-COV-2 variant calling was done using 3 different variant calling platforms, namely, CoVDB(*47*), Scorpio call v1.2.123 (https://pangolin.cog-uk.io/), and Nextclade v.1.13.2 (https://clades.nextstrain.org/)(49).

### Single nucleotide variation, and genetic diversity analysis

For assessing intrahost single nucleotide variation (iSNV), data from only those participants were included for whom sequence data from baseline and at least one follow-up time point were available. SNV analysis was performed using PASeq SARS-CoV-2 pipeline (www.paseq.org). Briefly, raw sequence files were quality filtered and adapter-trimmed using trimmomatic (v0.30). Contaminating sequences were filtered out using BBMap Suite (v35.76). Duplicated reads were detected using fastuniq (v1.1). High quality non-redundant reads were then aligned to SARS-CoV-2 Wuhan reference (NC_045512.2) using Bowtie2 (v.2.3.2). Resulting alignments were processed with samtools (v.1.2) and iVar (v1.4.2) to obtain nucleotide variant VCF files. Nucleotide variants present at 100% frequency of the total viral population at all time points indicative of lineage defining mutations were excluded from the iSNV analysis. Genetic diversity between multiple sequences of an individual were assessed by average pairwise distance in MEGA both at the nucleotide and amino-acid level.

### Statistical Analysis

Categorical variables were summarized using total number and percentage and between-group differences were evaluated using either chi-squared test or Fisher’s exact test when appropriate. Continuous variables were summarized with median and interquartile ranges and compared with non-parametric methods (Wilcoxon rank-sum test to compare two groups and Dunn’s test with Benjamini-Hochberg adjustment to compare three or more groups). Within group comparison was conducted using paired Wilcoxon signed-rank test without adjustment for multiple comparisons. We also used generalized estimating equation (GEE) with Gaussian estimation to evaluate between-group differences accounting for repeated measurement during longitudinal follow-up. R (4.3.0) was used for statistical analyses. Two-tailed tests were used for all the analyses and P<0.05 was considered statistically significant unless specified otherwise.

## Data Availability

All data produced in the present study are available upon reasonable request to the authors

## Acknowledgements

We thank all the participants for their participation in this important study. We thank all the healthcare workers who take care of them and kindly refer them to the POSITIVES study. This work was supported by the National Institutes of Health (U19 AI110818), the Massachusetts Consortium for Pathogen Readiness SARS-CoV-2 Variants Program and the Massachusetts General Hospital Department of Medicine. Drs. JA Sparks and ZS Wallace are supported by the National Institute of Arthritis and Musculoskeletal and Skin Diseases (R01 AR080659). Dr. JA Sparks is also supported by the Llura Gund Award funded by the Gordon and Llura Gund Foundation. Dr. GD Gaiha is supported by the NIH (DP2AI154421, R01AI176533 and DP1DA058476), the Bill and Melinda Gates Foundation, Burroughs Wellcome Career Award for Medical Scientists and Howard Goodman Fellowship. Dr. JZ Li was also supported by a grant from Merck. Dr. Y Li was supported by Rustbelt CFAR (Case Western Reserve University/University Hospitals Cleveland Medical Center and University of Pittsburgh, P30 AI036219). The BSL3 laboratory where viral culture work was performed is supported by the Harvard CFAR (P30 AI060354). The funders had no role in the study design; in the collection, analysis, and interpretation of data; in the writing of the manuscript; or in the decision to submit the manuscript for publication.

## Authors Contribution

Study conceptualization: MJ Siedner, AK Barczak, JE Lemieux, JZ Li

First draft writing: Y Li, MC Choudhary, J Regan, JZ Li

Draft review, discussion and approval: All authors

Statistical analyses: Y Li, MC Choudhary, R Deo, J Regan

Chart review and participant classification: Y Li, Y Kawano, ZS Wallace, JA Sparks

Virology assays: J Regan, J Boucau, GE Edelstein, C Marino, JP Flynn

Sequencing analyses: MC Choudhary, R Uddin, R Deo, JE Lemieux, JZ Li

Neutralizing antibody assays: T Speidel, MS Seaman Binding antibody assays: MY Liew

T cell assays: A Nathan, MA Getz, GD Gaiha

Study coordination, sample collection, participant referral: Z Reynold, M Barry, RF Gilbert, D Tien, S Sagar, TD Vyas, SP Hammond, LA Novack, B Choi, M Cernadas, ZS Wallace, JA Sparks, JM Vyas

Database management: MJ Siedner, RF Gilbert

## Supplementary Materials

## Supplementary Methods

### T cell proliferation staining panels

For PBMCs stained with carboxyfluorescein succinimidyl ester (CFSE; Life Technologies), cells were then washed and stained with anti-CD3 PE-Cy7 (clone SK7; BioLegend), anti-CD8 APC (clone SK1; BioLegend), anti-CD4 BV711 (clone RPA-T4; BioLegend), and LIVE/DEAD violet viability dye (Life Technologies).

For PBMCs stained with Cell Trace Far Red Proliferation Dye (CTFR, Invitrogen), cells were then washed and stained with anti-CD3 APC-Cy7 (clone UCHT1; BioLegend), anti-CD8 BV605 (clone SK1; BioLegend), anti-CD4 PE-Cy7 (clone OKT4; BioLegend), and LIVE/DEAD violet viability dye (Life Technologies).

### Generalized estimating equation (GEE)

GEE was performed using “geepack” package (version 1.3.9) in R^1^. In the GEE model, family was set as “gaussian”, and the correlation structure (“corstr”) was set as “independence”. Quasi Information Criterion (QIC) was used to compare models using “independence”, “exchangeable” and “ar1” and the one with “independence” had the lowest QIC. Monoclonal antibody (Mab) use, weeks since symptom onset or first positive PCR, numbers of vaccinations before enrollment, sex, and age were adjusted for in these models. Logarithm base 10 of the neutralizing antibody levels were treated as dependent variables and other variables as independent. Coefficients for all the independent variables were then transformed to the power of 10 and was shown in this figure as fold-change compared to reference group.

## Supplementary Tables

**Supplementary Table 1.** Categorization for immunocompromising conditions.

|  |  |  |
| --- | --- | --- |
| Non-Severe (NS) |  | • Autoimmune disease, receiving immunosuppressants that are not B cell/plasma cell targeted therapy within 12 months of study entry<br>• Solid malignant tumor on treatment (excluding those who underwent resection and were considered in remission after resection)<br>• Corticosteroid use equivalent to Prednisone >20mg daily for at least 14 consecutive days within 30 days prior to study entry<br>• HIV infection with CD4 cell count >200 cells/mm<sup>3</sup> |
| Severe | Severe- Hematological malignancy/Transplant (S-HT) | • Solid organ transplant (SOT)<br>• Hematopoietic stem cell transplant (HSCT)<br>• Lymphoma, leukemia<br>• Immune-Related Adverse Event (irAE) on multiple immunosuppressants targeting different pathways |
|  | Severe- Autoimmune and other B cell deficiency (S-A) | • Autoimmune disease receiving B cell targeted therapy within 12 months of study entry<br>• Congenital or late onset B cell deficiency (e.g. Common Variable Immunodeficiency) |

**Supplementary Table 2.** Diagnoses for immunocompromised participants and categorization.

| ID | Diagnosis | Immunosuppressants/Treatment | Group |
| --- | --- | --- | --- |
| 100 | Antiphospholipid syndrome | Prednisone, eculizumab, rituximab, cyclophosphamide | S-A |
| 101 | Heart and kidney transplant | Everolimus, Prednisone 5mg daily, Tacrolimus | S-HT |
| 107 | Rheumatoid arthritis | Tocilizumab | NS |
| 113 | Minimal change disease | Rituximab within 12 months of COVID-19 | S-A |
| 126 | RA | Abatacept+ Prednisone 5mg daily | NS |
| 217 | RA | Methotrexate+ Prednisone 5mg daily | NS |
| 240 | Chronic myelogenous leukemia | Dasatinib | S-HT |
| 245 | Diffuse large B-cell lymphoma | Rituximab+Polatuzumab+Prednisone, CAR-T, Chemotherapy, Tocilizumab | S-HT |
| 388 | Adenocarcinoma of pancreas | Chemotherapy and radiation | NS |
| 449 | Granulomatosis with polyangiitis | Rituximab within 12 months of COVID-19 | S-A |
| 459 | Immune Related Adverse Events, Metastatic Merkel cell carcinoma | Pembrolizumab, Tacrolimus, Prednisone, Mycophenolate | S-HT |
| 470 | Multiple sclerosis | Ocrelizumab within 12 months of COVID-19 | S-A |
| 471 | Marginal zone lymphoma (CR) | Obinutuzumab+CHOP | S-HT |
| 475 | Multiple sclerosis | Rituximab within 12 months of COVID-19 | S-A |
| 497 | Diffuse large B-cell lymphoma (CR), Hypogammaglobulinemia | Rituximab-CHOP | S-HT |
| 531 | Sarcoidosis | Infliximab | NS |
| 532 | Granulomatosis with polyangiitis | Mycophenolate | NS |
| 533 | Diffuse large B cell lymphoma, Hypogammaglobulinemia after CAR-T | CAR-T, Tocilizumab, Pembrolizumab, Rituximab, Corticosteroid | S-HT |
| 534 | RA, Sjogren's syndrome, Hypogammaglobulinemia; cryoglobulinemia | IVIG | S-A |
| 547 | RA | Tocilizumab+Methotrexate | NS |
| 548 | RA | Rituximab within 12 months of COVID-19+ Leflunomide | S-A |
| 549 | Bechet's disease | Azathioprine | NS |
| 550 | RA | Methotrexate+ Hydroxychloroquine | NS |
| 551 | Psoriatic arthritis | Infliximab | NS |
| 552 | Seronegative spondyloarthropathy | Adalimumab+ Methotrexate | NS |
| 557 | RA, SLE | Methotrexate | NS |
| 558 | SLE | Belimumab | S-A |
| 563 | RA | Adalimumab | NS |
| 564 | Multiple myeloma | Daratumumab+ Dexamethasone | S-HT |
| 569 | SLE | Hydroxychloroquine+ Methylprednisolone daily 6mg | NS |
| 573 | Inflammatory arthritis | Adalimumab+ Hydroxychloroquine | NS |
| 597 | Giant cell arteritis, polymyalgia rheumatica | Tocilizumab+ Prednisone orally 5mg daily | NS |
| 604 | CVID | IVIG every 4 weeks | S-A |
| 610 | Ulcerative colitis | Infliximab | NS |
| 658 | RA | Tocilizumab | NS |
| 678 | RA | Tofacitinib | NS |
| 687 | SLE, RA | Hydroxychloroquine, methotrexate | NS |
| 688 | Mantle cell lymphoma | Zanubrutinib+Venetoclax; Last dose of Obinutuzumab within 12 months | S-HT |
| 691 | RA | Rituximab | S-A |
| 708 | Minimal change disease | Rituximab | S-A |
| 716 | RA | Methotrexate | NS |
| 723 | Multiple sclerosis, acquired hypogammaglobulinemia | IVIG every 4 weeks; Ocrelizumab within 12 months | S-A |
| 725 | RA | Infliximab, methotrexate, hydroxychloroquine | NS |
| 735 | Psoriatic arthritis | Adalimumab | NS |
| 768 | Ankylosing spondylitis | Secukinumab | NS |
| 793 | RA | Adalimumab, methotrexate | NS |
| 805 | Ulcerative colitis, inflammatory arthritis | Golimumab, methotrexate | NS |
| 870 | Mantle cell lymphoma | Bendamustine and rituximab within 12 months | S-HT |
| 892 | HIV infection | N/A, on antiretroviral therapy, CD4 cell count>200 | NS |
| 936 | RA | Etanercept, hydroxychloroquine | NS |
| 945 | IgG4 related disease | Rituximab | S-A |
| 946 | Diffuse large B-cell lymphoma | R-CHOP | S-HT |
| 952 | Inflammatory arthritis | Adalimumab | NS |
| 953 | RA | Infliximab, methotrexate, prednisone 3mg | NS |
| 965 | Breast cancer | Trastuzumab deruxtecan within 12 months, then Olaparib | NS |
| 982 | Follicular lymphoma | Bendamustine, Obinutuzumab within 12 months | S-HT |

S-HT, severe hematological oncology/Transplant; S-A, severe autoimmune disease/B-cell deficient; NS, non-severe immunocompromise; CAR-T, chimeric antigen receptor T-cell therapy; CR, complete remission; CHOP, cyclophosphamide, doxorubicin, vincristine, and prednisone; R-CHOP, Rituximab in combination with CHOP; RA, rheumatoid arthritis; SLE, systemic lupus erythematosus; CVID, common variable immunodeficiency

**Supplementary Table 3.** Monoclonal antibody use.

|  | S-HT<br>(N=12) | S-A<br>(N=13) | NS<br>(N=31) | None<br>(N=184) | Overall<br>(N=240) |
| --- | --- | --- | --- | --- | --- |
| Bamlanivimab-Etesevimab | 1 | 0 | 0 | 4 | 5 |
| Casirivimab-Imdevimab | 0 | 2<sup>b</sup> | 2 | 5 | 9 |
| Sotrovimab | 4<sup>a</sup> | 1 | 1 | 0 | 6 |
| Bebtelovimab | 3 | 2 | 2 | 1 | 8 |
| Tixagevimab-Cilgavimab | 5<sup>c</sup> | 4 | 0 | 0 | 9 |

a, three participants received Sotrovimab after blood draws.
b, one participant received Casirivimab-Imdevimab after blood draws.
c, two participants received both Tixagevimab-Cilgavimab and Sotrovimab and one participant received both Tixagevimab-Cilgavimab and Bebtelovimab

## Supplementary Figures

**Supplementary Figure 1.**
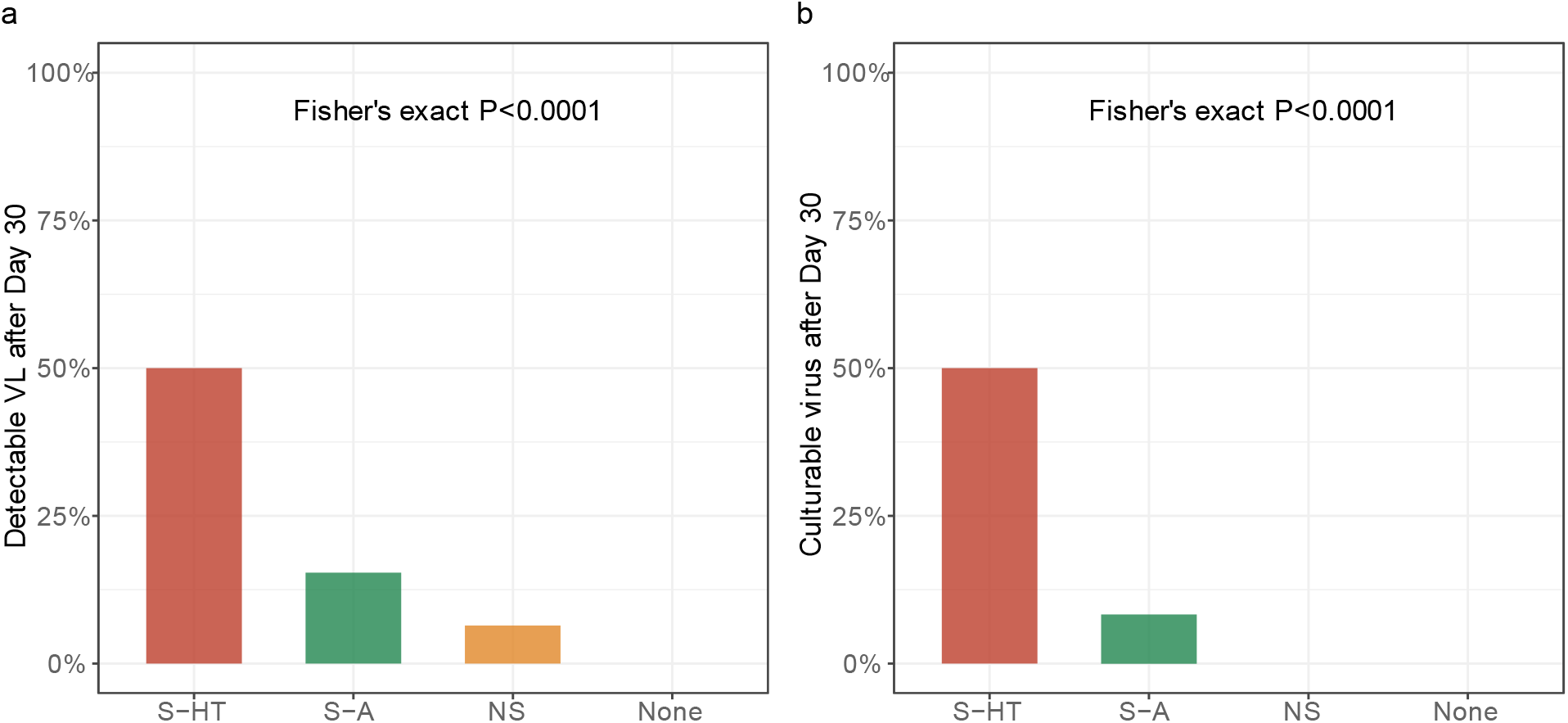
Detectable SARS-CoV-2 viral RNA (a) and culturable SARS-CoV-2 virus (b) beyond 30 days after symptom onset or first positive PCR/antigen tests, supplemental to Fig. 1. Fisher’s exact test was used to calculate the P values.

**Supplementary Figure 2.**
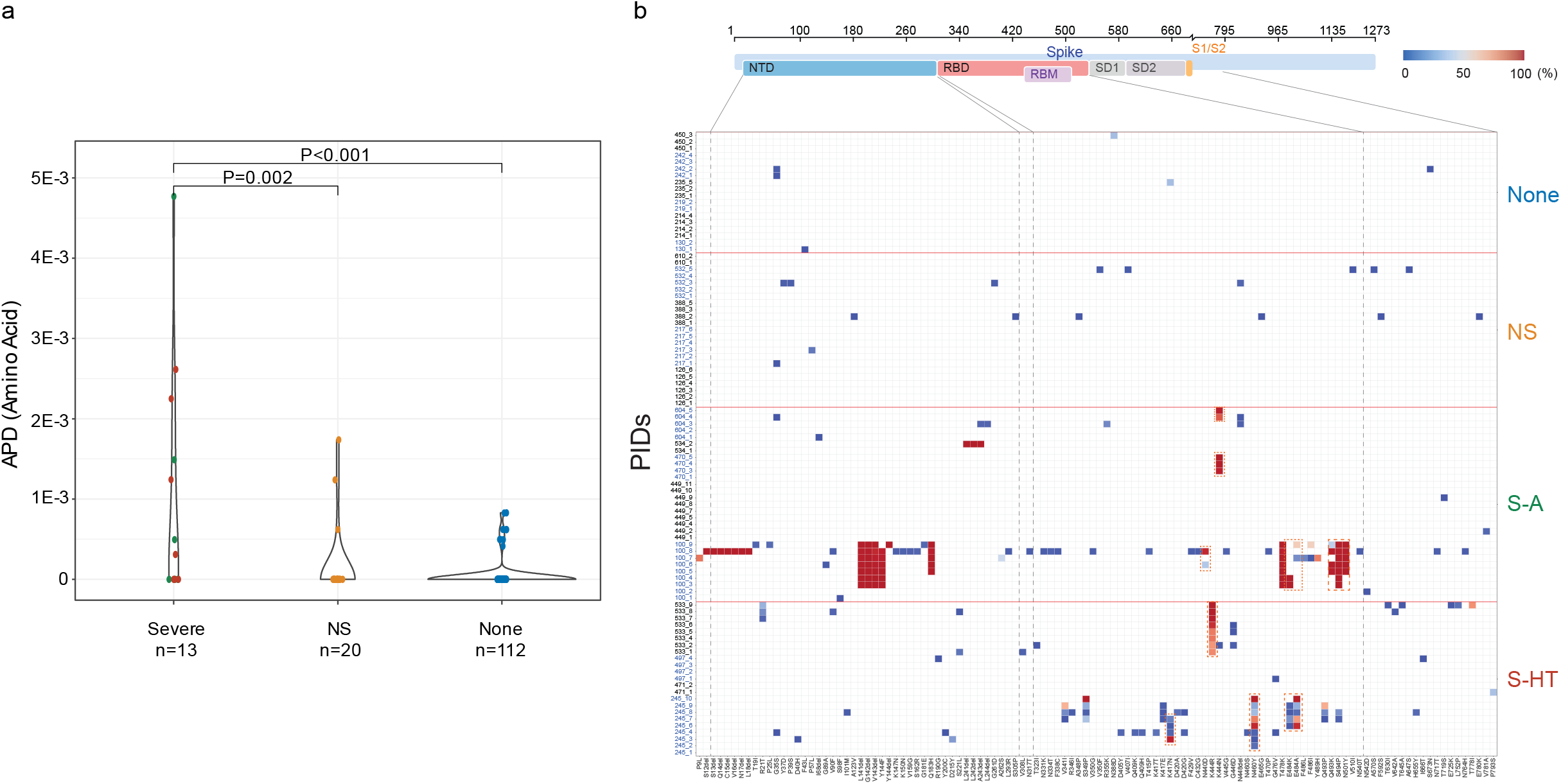
SARS-CoV-2 mutations among different immunocompromised groups, supplementary to Fig. 2. (a), SARS-CoV-2 intrahost mutations at the amino acid level among different immunocompromise groups. (b), Heat map showing distribution of Spike polymorphisms from participants receiving mAb treatment longitudinally. In the heatmap, y axis indicates participants’ ID (PID) followed by sequential numbers of sample collection, while x axis shows amino acid positions in the Spike gene. Different domains of Spike are shown at the top. Colors indicate frequency of polymorphisms, with blue indicating the lowest value and red indicating the highest value in the scale. Participants in different study groups are separated by a red horizontal line. mAb resistance mutations are shown by red dotted box.

**Supplementary Figure 3.**
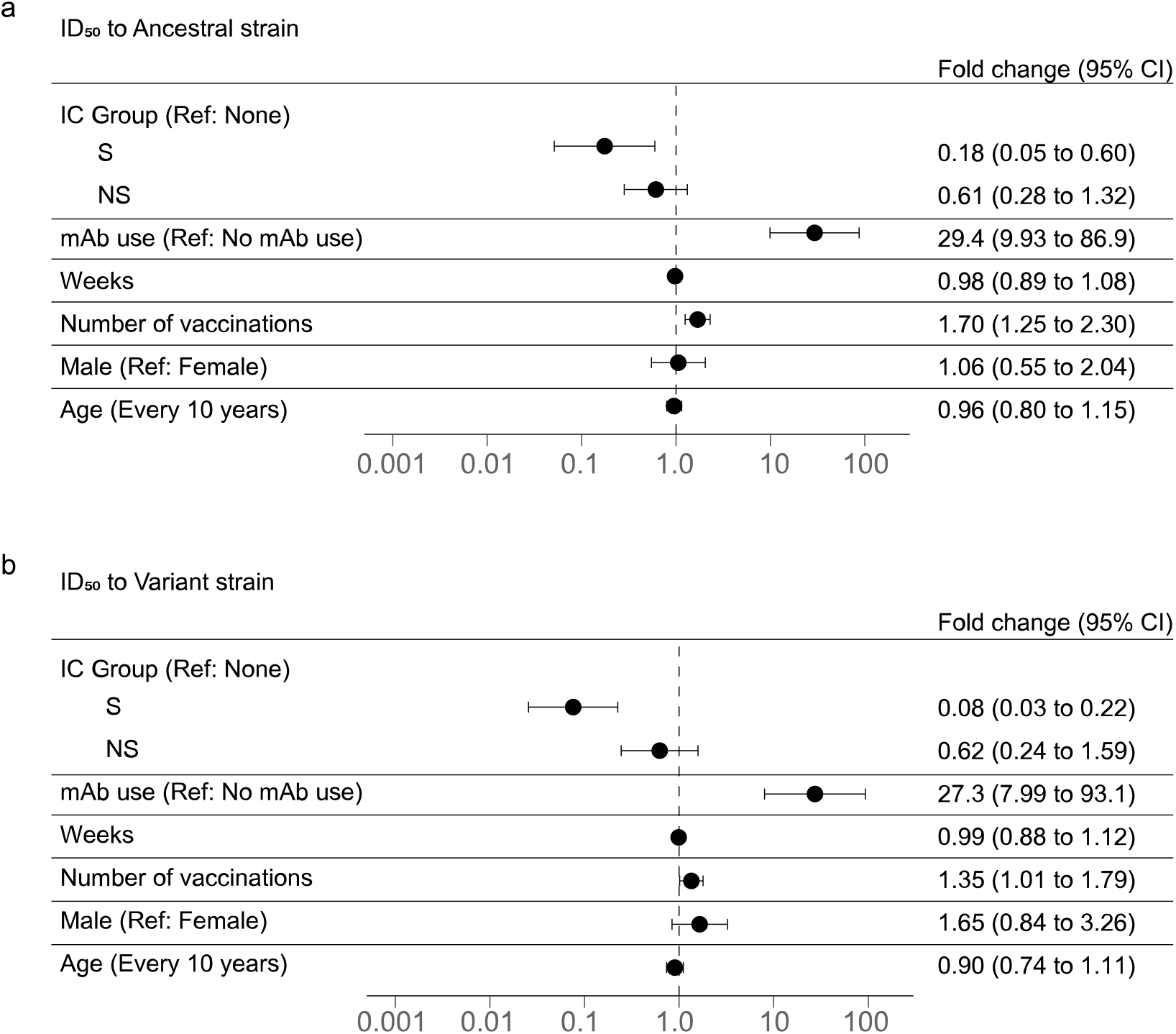
Severe immunocompromise is associated with lower neutralizing antibody levels, supplemental to Fig. 3. Generalized estimation equation to account for longitudinal repeated measurements was used to estimate the association between immunocompromise groups and neutralizing antibody levels against ancestral SARS-CoV-2 Spike protein (a) and variant-specific Spike protein (b). Monoclonal antibody (mAb) use, weeks since symptom onset or first positive PCR/antigen, numbers of vaccinations before enrollment, sex, and age were adjusted for in these models.

**Supplementary Figure 4.**
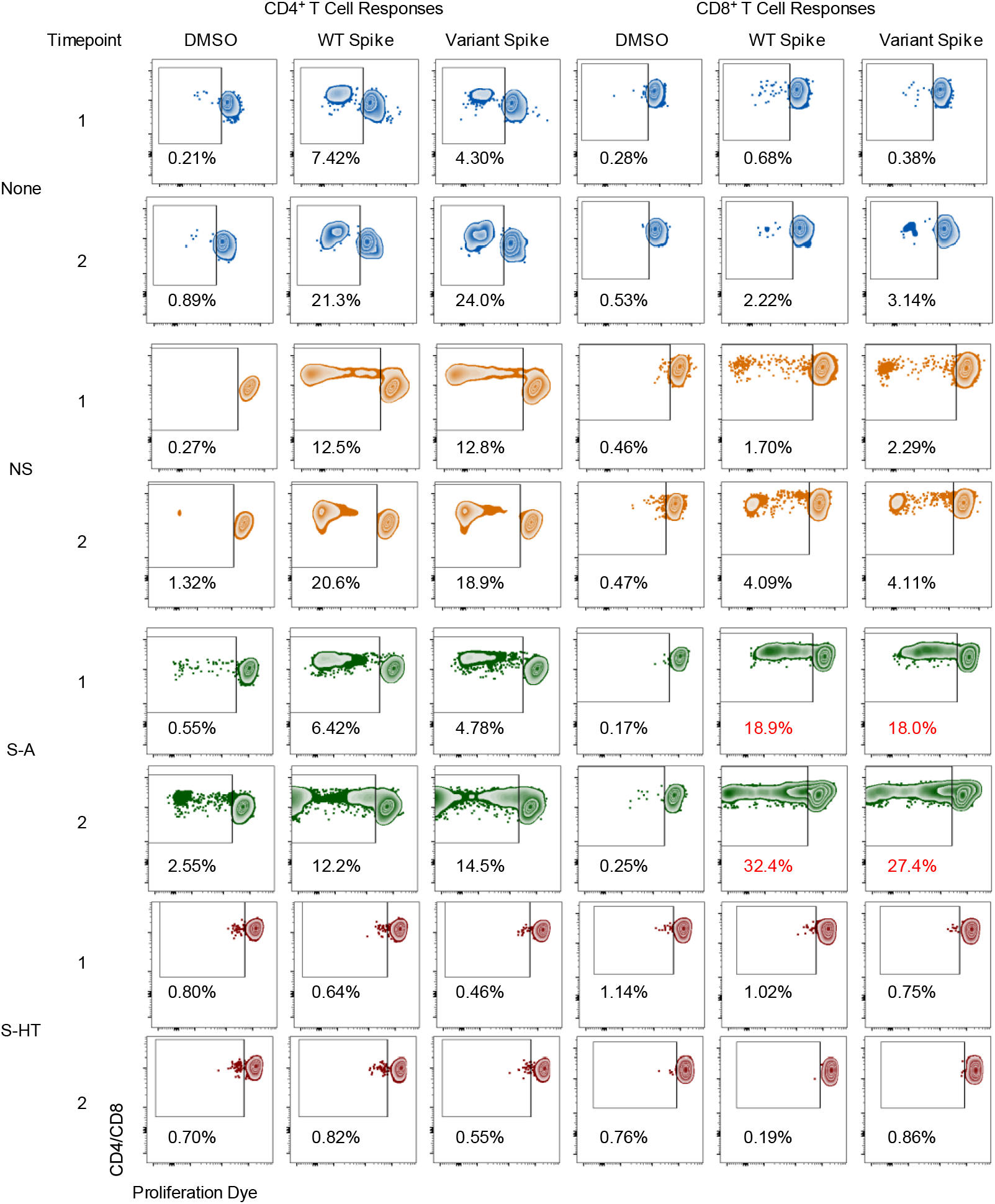
Representative T cell proliferation assay gating scheme, supplementary to Fig. 4. CD4+ and CD8+ T cell proliferation results from representative participants in each immunocompromise group are shown. Non-immunocompromised group, ID=261 (Omicron, BA.1); Non-severe group (NS), ID= 768 (Omicron, BF.5); Severe-autoimmune/B-cell deficient (S-A), ID=534 (Omicron, BA.2); Severe- hematological malignancy/transplant (S-HT), ID= 245 (Delta, B.1.617.2).

**Supplementary Figure 5.**
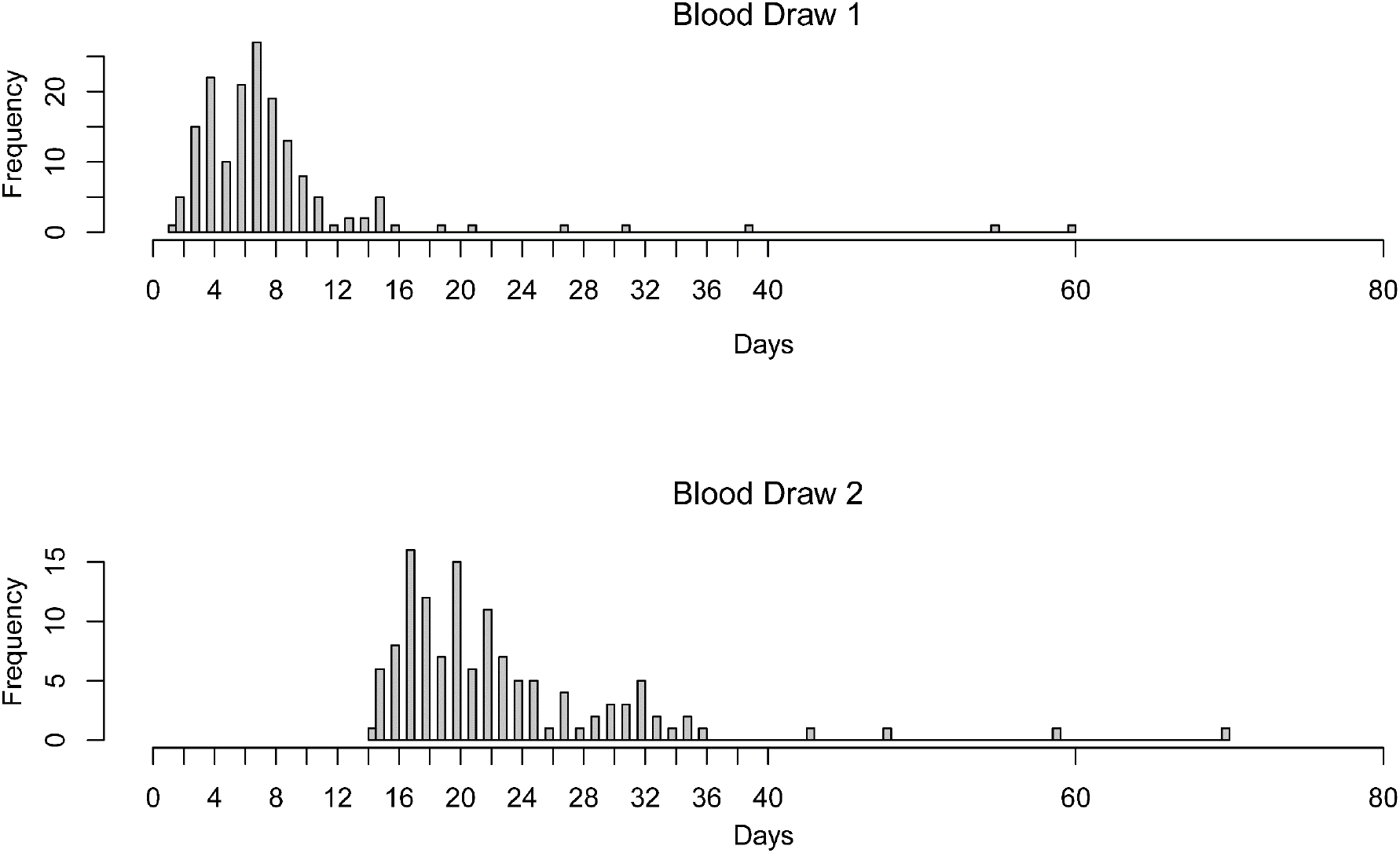
Distribution of duration between symptom onset or first positive PCR and blood draws.

